# Symptom burden, Psychosocial Distress, Resilience and Health-Related Quality of Life in Women with Metastatic Breast Cancer Receiving Contemporary Systemic Therapy

**DOI:** 10.64898/2026.09.08.26362524

**Authors:** Yan Zhan, Shelli Feder, Sangchoon Jeon, Maryam Lustberg, Q. Margaret Rosenzweig, Djin Tay, M. Tish Knobf

## Abstract

**Aims:** Targeted therapies and immunotherapies have extended survival of women with metastatic breast cancer. However, the role of resilience among women receiving those contemporary treatments remains understudied. We aimed to examine the interrelationships among physical symptom burden, psychosocial distress, resilience and health-related quality of life in women with metastatic breast cancer receiving contemporary systemic cancer treatment.

**Design:** Cross-sectional study.

**Methods:** Women with metastatic breast cancer receiving systemic treatment were recruited across the United States from February to September 2024. Data was collected using validated measures. Psychosocial distress was constructed as a latent variable indicated by anxiety, depression and uncertainty, using confirmatory factor analysis. We conducted structural equation modeling to examine multiple pathways between physical symptom burden, latent psychosocial distress, resilience and health-related quality of life.

**Results:** Of 217 participants, 209 were included in the analysis (mean age = 50.1 years, SD=14.1). Physical symptom burden, psychosocial distress and resilience were significantly associated with health-related quality of life. Significant indirect associations of physical symptom burden and psychosocial distress with health-related quality of life through resilience were identified. Structural equation modeling showed indirect associations between physical symptom burden and health-related quality of life through psychosocial distress and resilience (*β* =-0.137, 95% CI: -0.217, -0.078, *p*<0.001).

**Conclusion:** Psychosocial distress appeared to be more proximally associated with resilience than physical symptom burden among this population. Resilience interventions may be strengthened by identifying and addressing anxiety, depression and uncertainty as proximal intervention points for preserving resilience, rather than only including generic stress management components.

**Impact:** This study highlighted the need of resilience interventions that prioritize anxiety and depression screening, mental health referral, and distress focused supportive care alongside symptom management for this population.

**Reporting Method:** STROBE, Cross-sectional guidelines.

**Patient or Public Contribution:** This study involved women with metastatic breast cancer and breast cancer patient advocacy groups participated in recruitment.

## 1. Introduction

Women with metastatic breast cancer (MBC) account for 30-40 % of breast cancer survivors [1], among them 5-10% were diagnosed denovo (present at diagnosis) and up to 30% of those initially diagnosed with early-stage (I-II) breast cancer eventually develop metastatic disease (stage IV). Advances in contemporary systemic cancer treatments, such as targeted therapies and immunotherapies, have extended survival for many women with MBC [2, 3]. As a result, MBC can be managed as a chronic, life-limiting condition in which women may live for prolonged period of time while receiving sequential lines of chemotherapy with targeted or immunotherapies [52]. However, longer survivorship also means extended exposure to treatment-related toxicities, cancer-related symptoms, and persistent psychosocial stressors [4–6]. Despite therapeutic advancement, how contemporary systemic treatment contribute to physical symptom burden, psychosocial well-being and health-related quality of life (HRQOL) of women with MBC remains understudied.

HRQOL is a multidimensional construct encompassing physical, psychological, social and functional well-being [7, 8], and it served as an important clinical indicator for people with metastatic cancer [9]. Women with MBC historically reported low HRQOL and high symptom burden, including pain, fatigue, nausea, and sleep disturbance [10, 11]. These physical symptoms that occur in the context of living with incurable cancer can contribute to psychological distress. Prior studies have shown that women with MBC experienced high level of anxiety and depressive symptoms [6,12], and uncertainty [13,14] related to disease progression, treatment responses, and future prognosis. The use of contemporary systemic treatments may sustain patients’ hope to live longer but can also introduce complex and unpredictable treatment-related toxicities and side effects, which require patients to continually adapt to changing physical and emotional demands. In a recent literature review, physical symptom burden and psychosocial distress remain the major stressors that adversely impact HRQOL among women with MBC [15], but majority research evidence reviewed has not examined contemporary systemic treatment, particularly targeted therapies and immunotherapies.

Resilience is defined as an ability to “bounce back” including the process of adaptation and adjustment to external or internal stressors [16,17, 18]. Resilience theories, including the Kumpf’s resilience framework [19] and resilience framework for nursing and healthcare [20], described stressors and adversities (e.g., chronic illness, trauma), and the resilience process through which individuals respond to adversities by utilizing various resources and engaging protective, compensatory and coping strategies to achieve functional adaptation. In cancer population, higher resilience has been associated with improved HRQOL and overall well-being [21]. Patients with greater resilience showed higher optimism, self-efficacy, and proactive coping, which support adjustment of patients during cancer trajectory [22]. Previous studies have suggested that resilience mediated the relationships between symptoms and HRQOL [23, 24], and between depression and health status [51], particularly among patients with early-stage cancer or chronic illnesses. However, whether and how resilience is associated with symptom burden, psychosocial distress, and HRQOL among women with MBC receiving contemporary systemic treatments has not been well studied.

This gap is important because supportive care remained scarce for women with MBC to support and rebuild their lives after cancer treatment. There is mounting evidence that physical symptoms provoke negative psychosocial responses and contribute to poorer HRQOL [12, 25, 26]. Compared with patients with early-stage cancers, women with MBC experience longer treatment regimens, more episodes of treatment response and resistance, disease progression and uncertainty while seeking benefit from life-prolonging therapies. Although prior studies have established associations among symptom burden, psychosocial response, resilience and HRQOL, less is known about how these factors are interrelated in women with MBC in the contemporary treatment era, as this population has been predominantly excluded from prior research. Clarifying these relationships may help to identify modifiable supportive care targets to improve HRQOL in this understudied population.

## 2. The Study

The purpose of this study was to examine interrelationships among physical symptom burden, psychosocial distress, resilience and HRQOL, as well as the role of resilience in women with MBC. We proposed that resilience mediates the association between physical symptom burden, psychosocial distress and HRQOL; and physical symptom burden indirectly associated with HRQOL through psychosocial distress and resilience among women with MBC.

## 3. Methods

### 3.1 Study design

We employed a descriptive cross-sectional design and administrated patient-reported questionnaires to collect data from women with MBC undergoing contemporary systemic cancer treatment.

### 3.2 Inclusion and Exclusion Criteria

Inclusion criteria included 1) women diagnosed with MBC; 2) currently on systemic cancer treatment (i.e., chemotherapy, hormonal therapy, targeted therapy and immunotherapy) and 3) were literate in English. Women who were unable to consent or had a second cancer diagnosis at the time of recruitment were ineligible to the study.

### 3.3 Study Sampling and Setting

We conducted a power analysis for the sample size based on the effect that detect partial correlations of 0.273 (=0.39*0.7) to 0.455 (=0.65*0.7) for the primary predictors (cancer symptoms, psychosocial distress, resilience) on HRQOL by assuming a 30% reduction after adjusting for covariates. These effect sizes were found in the prior study [27]. We estimated the sample size of 200 will have 90% power in multiple regression with covariates at a 5% significance level. For mediation analysis, we estimated adjusted mediation effects of -0.156 to - 0.135 by assuming a medium-size coefficient of -0.30 for main predictors. We performed a power analysis using the Monte Carlo Power Analysis tool to detect medication effects [28]. With a sample size of 200, we can detect the mediation effect size with approximately 95% power.

Using the convenience sampling method, we recruited 217 women participants from a comprehensive cancer center in the U.S, and five MBC patient advocacy groups across the country. Participants were recruited from February to September 2024. Among 217 participants, 209 completed the questionnaires, yielding a completion rate of 96.3%. This sample size had enough statistical power and thus was appropriate for this study. The detailed study procedures and data collection have been described in the previous study published elsewhere [50].

### 3.4 Measures

#### 3.4.1 Physical symptom burden

Physical symptom burden was measured by the MD Anderson Symptom Inventory (MDASI), including 13 core symptoms (e.g., pain, fatigue, disturbed sleep, etc.) [29]. Participants rated the severity of each symptom over the past week on a 0-10 rating scale, with a higher total score indicating greater symptom burden. Cronbach’s α for MDASI in this study was 0.93.

#### 3.4.2 Psychosocial distress

Psychosocial distress was defined as adverse psychosocial outcomes, which indicated by anxiety, depression, and uncertainty symptoms. Anxiety and depression were assessed using the 6-item Patient-Reported Outcomes Measurement Information System (PROMIS) Anxiety short form 6a [30] and PROMIS Depression short form 6a [31], respectively. Each item was rated on a 5-point Likert scale (1-“Never” to 5-“Always”), with a higher score indicating greater anxiety and depression. Raw scores were converted into T-scores according to the PROMIS scoring manuals [32]. Uncertainty was measured using a 5-item Short Form of the Mishel Uncertainty in Illness Scale (SF-MUIS) [33]. Items are rated from 1= “strongly disagree” to 4= “strongly agree.” The positive statement items were reverse-coded. Total scores range from 5 (no illness uncertainty) to 25 (high illness uncertainty).

#### 3.4.3 Resilience

Resilience was measured by the 10-item Resilience Scale Specific to Cancer (RS-SC-10). The instrument was developed and validated by Ye and colleagues [34], which contained 10 items that capture resilience specifically related to cancer. The instrument included a five-point Likert scale ranging from 1= “Never” to 5= “Always”. The total score ranges from 10 to 50, with higher scores indicating higher levels of resilience. The RS-SC-10 has been validated in US breast cancer population, with a reported Cronbach’s α of 0.86 [35]. Cronbach’s α for RS-SC-10 in this study was 0.79. In this study, resilience served as a mediator.

#### 3.4.4 HRQOL

HRQOL was measured by the Functional Assessment of Cancer Therapy-General (FACT-G) [36], a 27-item self-report instrument that assesses HRQOL among cancer patients. Items were rated on a 5-point Likert scale ranging from 0= “Not at All” to 4=“Very Much.” A total FACT-G score was calculated by summing all items, with higher total scores reflecting higher HRQOL [36].

#### 3.4.5 Sociodemographic and Clinical Characteristics

The sociodemographic characteristics (e.g., age, race, ethnicity, educational level, and marital status, etc.) and clinical factors (e.g., MBC subtype, current breast cancer treatment, time on the current treatment) were selected as they were associated with resilience, and physical symptoms, psychosocial distress and health-related quality of life in literature.

### 3.5 Statistical Analysis

We used R statistical software (version 4.4.1) [37] to conduct data analyses. Descriptive statistics, including frequency, percentages, means, standard deviations and ranges, were conducted to summarize sociodemographic and clinical characteristics. One-way ANOVA was employed to examine differences in HRQOL (FACT-G) across categorical variables. We conducted Pearson and Spearman correlation coefficients to assess the associations between continuous variables and the FACT-G score. Bivariate correlations were then examined among physical symptom burden, psychosocial distress (anxiety, depression, uncertainty, resilience), and HRQOL.

To evaluate the associations of physical symptom burden, psychosocial distress (anxiety, depression, and uncertainty), and resilience with HRQOL, we conducted multiple linear regression analyses adjusting for sociodemographic and clinical variables associated with HRQOL with p-values < 0.1. Model diagnostics were performed to assess outliers and evaluate assumptions of linearity, normality, and homoscedasticity using residual plots, the Shapiro-Wilk test and the Breusch-Pagan test, respectively. We also checked multicollinearity using the variance inflation factor (VIF). The values of VIF ranged from 1.29 to 2.84, indicating there was no multicollinearity.

We conducted structural equation modelling (SEM) to examine multiple pathways among physical symptom burden, psychosocial distress, resilience and HRQOL and the role of resilience. As anxiety, depression and uncertainty often occur at the same time when individuals appraised MBC disease as a life-threatening event, we thus combined all three psychosocial symptoms into one reliable latent variable. Confirmatory factor analysis (CFA) was used to evaluate the loadings of each psychosocial symptom and validated this latent psychosocial distress variable.

Three SEM models were conducted to compare and examine the multiple pathways among physical symptom burden, psychosocial distress, resilience and HRQOL. In Model 1, we examined whether resilience mediated the association between physical symptom burden and HRQOL. In Model 2, we assessed whether resilience mediated the association between psychosocial distress and HRQOL. In Model 3, we examined the psychosocial distress (first mediator) and resilience (second mediator) sequentially mediated association between physical symptom burden and HRQOL. All models were estimated using the full information maximum likelihood (FIML) to handle missing data. Standardized coefficient (β), standard errors, and 95% confidence intervals (CIs) were calculated for direct, indirect, and total effects.

We used the “Lavaan” package in R statistical software for CFA and SEM analyses. The significance of indirect effects was tested using bootstrapping with 5,000 resamples to obtain bias-corrected 95% CIs. Mediation was considered statistically significant if the 95% CI did not include zero, and p-value < 0.05. Covariates that were significant in bivariate analyses were included in SEM models. Model fit was evaluated using the following goodness-of-fit indices: Chi-square to degrees of freedom ratio (χ^2^/df ≤ 3), Root-mean-square error of approximation (RMSEA ≤ 0.06); Standard root mean residual (SRMR≤ 0.08), and Comparative Fit Index (CFI) and Tucker-Lewis Fit Index (TLI ≥ 0.95 indicating excellent fit) [38].

### 3.6 Ethical Considerations

This study received ethical approval from the Yale University Institutional Review Board (IRB ID: HIC# 2000036269). All individuals provided informed consent before participating in the study.

## 4. Results

### 4.1 Characteristics of Participants and bivariate analyses

Table 1 describes sociodemographic, clinical characteristics, and their bivariate associations with HRQOL. The mean age of the participants was 50.1 years (SD=14.1, range 24-85). Majority participants (76.6%) identified as White, over 85% were non-Hispanic, and nearly 80% had completed a college degree or above. The most common MBC subtype was HR+/HER2-(65.1%). The mean number of completed lines of treatment was 2.4 (range 0-8) and the mean time since MBC diagnosis was 4.4 years (SD=3.9).

**Table 1.** Bivariate Associations Between Sociodemographic, Clinical Characteristics and Health-related Quality of Life Score (FACT-G), N=209.

| Characteristics | FACT-G |  |  |  |
| --- | --- | --- | --- | --- |
|  | N (%) | Mean (SD) | r/F | P-value |
| Race |  |  | 2.77<sup>b</sup> | .019 |
| White | 160 (76.6) | 68.2 (17.9) |  |  |
| Black or African American | 18 (8.6) | 60.7 (15.3) |  |  |
| Other | 14 (6.7) | 58.9 (18.1) |  |  |
| American Indian or Alaska Native | 8 (3.8) | 52.5 (13.2) |  |  |
| Asian | 5 (2.4) | 60.4 (6.6) |  |  |
| Native Hawaiian or Other Pacific Islander | 2 (1) | 47 (39.6) |  |  |
| Missing | 2 (1) | - |  |  |
| Ethnicity |  |  | 5.15<sup>b</sup> | .006 |
| Non-Hispanic | 178 (85.2) | 67.4 (17.8) |  |  |
| Hispanic | 22 (10.5) | 57.7 (18.2) |  |  |
| Do not know | 7 (3.4) | 52.3 (6.6) |  |  |
| Missing | 2 (1) | - |  |  |
| Marital Status |  |  | 1.17<sup>b</sup> | .324 |
| Married/Cohabiting | 147 (70.3) | 66.6 (17.5) |  |  |
| Divorced/Separated | 33 (15.8) | 62.9 (19.1) |  |  |
| Never Married | 17 (8.1) | 61.6 (19.9) |  |  |
| Widowed | 10 (4.8) | 72.6 (17.7) |  |  |
| Missing | 2 (1) | - |  |  |
| Educational Level |  |  | 3.59<sup>b</sup> | .015 |
| High school graduate | 4 (1.9) | 67 (20.9) |  |  |
| Some college | 36 (17.2) | 60.5 (19.2) |  |  |
| College | 87 (41.6) | 63.6 (18.9) |  |  |
| Master's degree or above | 80 (38.3) | 70.7 (15.2) |  |  |
| Missing | 2 (1) | - |  |  |
| Employment Status |  |  | 9.96<sup>b</sup> | <.001 |
| Unemployed | 12 (5.7) | 55.2 (15.5) |  |  |
| Employed full-time | 69 (33) | 68.5 (16.6) |  |  |
| Employed part-time/self-employed | 33 (15.8) | 72.8 (13.7) |  |  |
| Retired | 38 (18.2) | 73.2 (17.5) |  |  |
| Disabled | 55 (26.3) | 55.9 (17.7) |  |  |
| Missing | 2 (1) | - |  |  |
| Household Annual Income |  |  | 4.84<sup>b</sup> | .003 |
| <\$50,000 | 38 (18.2) | 58.2 (19.2) | | |
| \$50,000 - \$100,000 | 71 (34) | 64.3 (17.3) | | |
| \$101,000 - \$150,000 | 50 (23.9) | 68.3 (18.2) | | |
| >\$150,000 | 48 (23) | 71.9 (15.6) | | |
| Missing | 2 (1) | - |  |  |
| Insurance |  |  | 5.39<sup>b</sup> | .001 |
| Medicare | 38 (18.2) | 59.23 (20.2) |  |  |
| Medicaid | 14 (6.7) | 56.6 (19.1) |  |  |
| Private insurance/Marketplace | 86 (41.2) | 70.7 (16.2) |  |  |
| Other | 69 (33) | 65.5 (16.9) |  |  |
| Missing | 2 (1) | - |  |  |
| Religion |  |  | 1.01<sup>b</sup> | .404 |
| Christian | 92 (44) | 63.9 (18.4) |  |  |
| Catholic | 33 (15.8) | 64.8 (18.7) |  |  |
| Muslim | 4 (1.9) | 60.3 (16.6) |  |  |
| Judaism | 11 (5.3) | 70.5 (14.4) |  |  |
| Other/none | 67 (31.6) | 68.7 (17.6) |  |  |
| Missing | 2 (1) | - |  |  |
| Dependent Children |  |  | 1.22<sup>b</sup> | .271 |
| Yes | 97 (46.4) | 64.4 (18.2) |  |  |
| No | 110 (52.6) | 67.2 (17.8) |  |  |
| Missing | 2 (1) | - |  |  |
| Hormone receptor status |  |  | 0.65<sup>b</sup> | .524 |
| Positive | 170 (81.3) | 66.2 (17.4) |  |  |
| Negative | 36 (17.2) | 66.4 (21.1) |  |  |
| Do not know | 3 (1.4) | 54.3 (4.0) |  |  |
| HER-2 status |  |  | 0.65<sup>b</sup> | .526 |
| Positive | 56 (26.8) | 66.3 (18.0) |  |  |
| Negative | 150 (71.8) | 66.2 (18.1) |  |  |
| Do not know | 3 (1.4) | 54.3 (4.0) |  |  |
| Tumor Characteristics |  |  | 0.36<sup>b</sup> | .838 |
| HR+/HER2- | 136 (65.1) | 66.3 (17.9) |  |  |
| HR+/HER2+ | 34 (16.3) | 65.6 (15.6) |  |  |
| HR-/HER2+ | 22 (10.5) | 67.2 (21.7) |  |  |
| HR-/HER2- | 14 (6.7) | 65.2 (20.8) |  |  |
| Do not know | 3 (1.4) | 54.3 (4.04) |  |  |
| Current treatment(s) |  |  | 2.92<sup>b</sup> | .035 |
| Chemo/chemo + targeted or immunotherapy | 79 (37.8) | 63.8 (17.6) |  |  |
| Hormone/hormonal +/-targeted or immunotherapy | 92 (44) | 66.3 (17.9) |  |  |
| Targeted alone | 34 (16.3) | 72.5 (17.8) |  |  |
| Do not know treatment Rx | 4 (1.9) | 50.8 (16.2) |  |  |
| Treatment(s) modalities |  |  | 1.04<sup>b</sup> | .388 |
| Oral pills | 61 (29.2) | 68.9 (16.0) |  |  |
| Intravenous (IV) treatment | 45 (21.5) | 67.1 (18.3) |  |  |
| Both oral and IV | 29 (13.9) | 61.1 (16.9) |  |  |
| Injection | 5 (2.4) | 65.8 (19.4) |  |  |
| More than one modality | 69 (33.3) | 64.9 (19.7) |  |  |
| Time on the current treatment(s) |  |  | 1.38<sup>b</sup> | .232 |
| < 1 year | 88 (42.1) | 67.7 (18.1) |  |  |
| 1 year | 37 (17.7) | 68.9 (16.9) |  |  |
| 2 years | 30 (14.4) | 60.8 (16.4) |  |  |
| 3 years | 21 (10.1) | 62.0 (20.1) |  |  |
| 4 years | 9 (4.3) | 59.7 (10.6) |  |  |
| More than 5 years | 24 (11.5) | 68.1 (20.4) |  |  |
| Palliative care enrollment |  |  | 11.75<sup>b</sup> | <.001 |
| Yes | 92 (44) | 61.4 (17.4) |  |  |
| No | 117 (56) | 69.7 (17.6) |  |  |
| Clinical trial(s) enrollment |  |  | 0.86<sup>b</sup> | .355 |
| Yes | 57 (27.3) | 64.2 (16.0) |  |  |
| No | 152 (72.7) | 66.8 (18.7) |  |  |
|  | Median (Range) | Mean (SD) |  |  |
| Age | 49 (24-85) | 50.1 (14.1) | 0.13<sup>a</sup> | .063 |
| Number of treatment line(s) completed | 2 (0-8) | 2.4 (1.8) | -0.26<sup>a</sup> | <.001 |
| Time since MBC diagnosis (in years) | 3 (0-19) | 4.4 (3.9) | -0.07<sup>a</sup> | .313 |

Note: SD= Standard deviation, a= r values of Spearman or Pearson correlation, b= F values of the analysis of variance test (ANOVA); participants with missing data were not analyzed as a separate category

In bivariate analyses, participants who identified as Black or African American, American Indian or Alaska Native, Asian and Native Hawaiian or other Pacific Islander, Hispanic, were unemployed, had household annual income <$50,000, or Medicaid insurance coverage reported lower HRQOL than their counterparts (Table 1). Additionally, participants enrolled in palliative care had lower FACT-score than those who did not (61.4±17.4 vs. 69.7±17.6, p<0.001). A higher number of completed treatment lines was significantly associated with lower HRQOL (r = -0.26, p<0.001).

### 4.2 Correlations of Variables and Multiple Linear Regression Analyses

Table 2 reports correlations among physical symptom burden, anxiety, depression and uncertainty, resilience and FACT-G score. The mean physical symptom burden (MDASI total score) was 3.86 (SD = 2.21), the average anxiety T-score was 58.85 (SD = 9.18), 55.40 (SD = 9.24) for depression, and the mean score was 11.92 (SD = 3.80) for uncertainty, 34.79 (SD = 5.96) for resilience and 66.05 (SD = 17.98) for FACT-G. Physical symptom burden, anxiety, depression and uncertainty were positively correlated with one another but negatively correlated with resilience and HRQOL (from r = -0.237 to -0.721, p<0.001 for all). Resilience was positively associated with HRQOL (r = 0.624, p<0.001). The results of unadjusted and adjusted multiple regression analyses with HRQOL are shown in Supplemental Table 1. After adjusting for sociodemographic and clinical covariates, physical symptom burden (β = -0.674, p<0.001), anxiety (β = -0.484, p<0.001), depression (β = -0.613, p<0.001) and uncertainty (β = -0.286, p<0.001) remained significantly and negatively associated with HRQOL.

**Table 2.** Correlation coefficients between Physical symptom burden, psychological distress, and HRQOL.

| **Coefficients** | **1** | **2** | **3** | **4** | **5** | **6** |
| --- | --- | --- | --- | --- | --- | --- |
| 1. Physical symptom | 1.000 |  |  |  |  |  |
| 2. Anxiety | 0.555** | 1.000 |  |  |  |  |
| 3. Depression | 0.614** | 0.723** | 1.000 |  |  |  |
| 4. Uncertainty | 0.428** | 0.351** | 0.447** | 1.000 |  |  |
| 5. Resilience | -0.335** | -0.380** | -0.523** | -0.237* | 1.000 |  |
| 6. FACT_G | -0.721** | -0.581** | -0.669** | -0.413** | 0.624** | 1.000 |
| Range | 0.08-10 | 39.1-82.4 | 38.4-80.2 | 5-24 | 20-50 | 17-106 |
| Mean (±SD) | 3.86 (2.21) | 58.85 (9.18) | 55.40 (9.24) | 11.92 (3.80) | 34.79 (5.96) | 66.05 (17.98) |

Note: \*\* P<.001; \* P<.01

### 4.3 Mediation Effects and Structural Equation Modeling

Table 3 summarizes the direct and indirect effects of Model 1 and 2. In Model 1 (Figure 1), resilience mediates the association between physical symptom burden and HRQOL. We found that physical symptom burden was negatively associated with resilience (β = -0.320, p<0.001), and resilience was associated with higher HRQOL (β = 0.453, p<0.001). The mediation effect of resilience on the relationship between physical symptom burden and HRQOL was statistically significant (Indirect effect: β = -0.145, 95% CI: -0.221, -0.072, p=0.001; Total effect: β = -0.655, 95% CI: -0.752, -0.552, p<0.001). Resilience accounted for 22.1% of the total effect of physical symptom burden on HRQOL.

**Figure 1.**
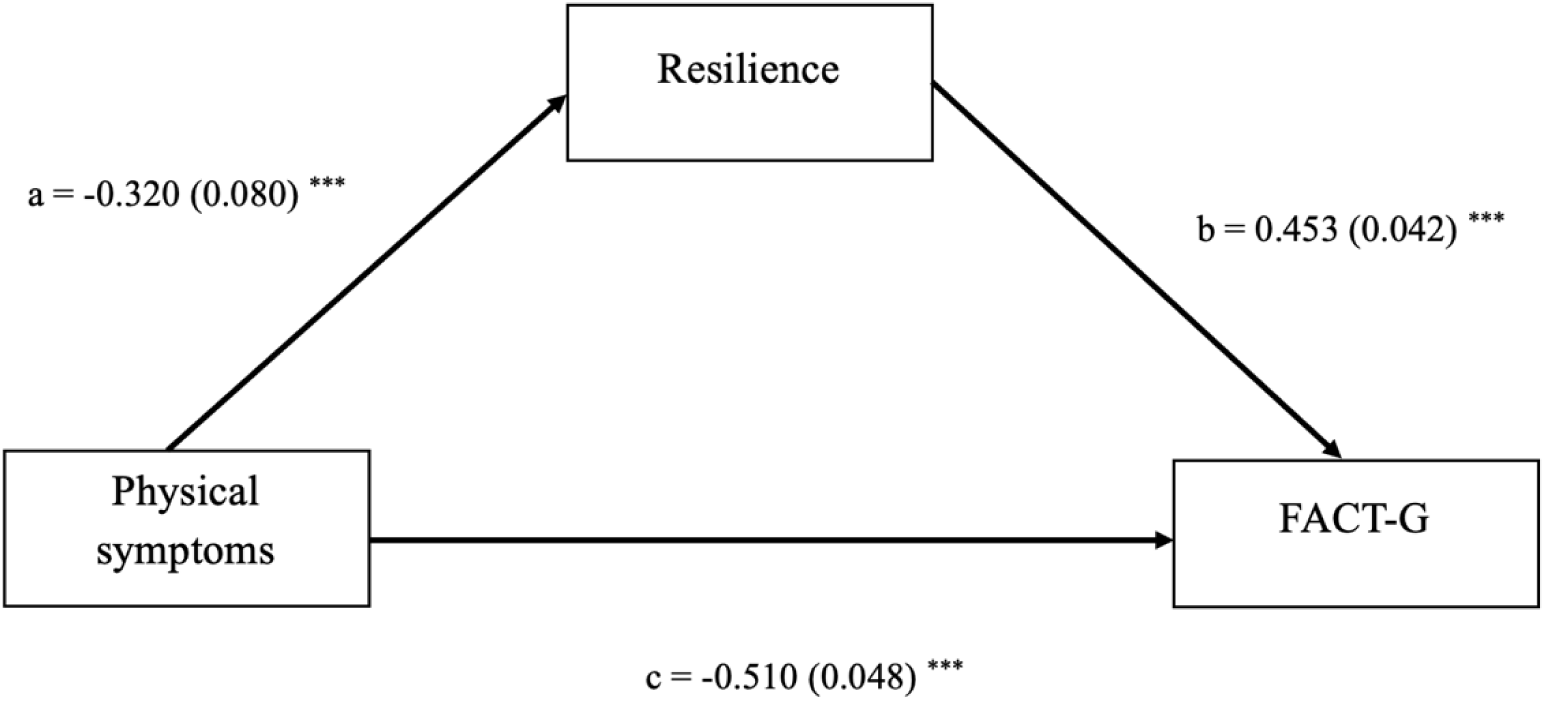
Mediation model of resilience in the relationship between physical symptom burden and HRQOL (FACT-G) Note: FACT-G = Health-related quality of life (HRQOL); a= effects of physical symptoms on resilience; b= effects of resilience on FACT-G adjusting for physical symptoms and other covariates; c= direct effects of physical symptoms on FACT-G adjusting for resilience and covariates. *** p<0.001

**Table 3.** Estimated Direct and Indirect (Mediation) Effects of Physical Symptom burden (Model 1) and Psychological Distress (Model 2) on HRQOL through Resilience.

|  | Model 1: Effects of Physical Symptom Burden on HRQOL |  |  |  | Model 2: Effects of Psychological Distress on HRQOL |  |  |  |
| --- | --- | --- | --- | --- | --- | --- | --- | --- |
| | $\beta$ | SE | 95% CI | P-values | $\beta$ | SE | 95% CI | P-values |
| Direct effect of predictor | -0.510 | 0.048 | -0.606, -0.414 | <0.001 | -0.521 | 0.066 | -0.642, -0.389 | <0.001 |
| Indirect effect of resilience | -0.145 | 0.038 | -0.221, -0.072 | 0.001 | -0.180 | 0.037 | -0.256, -0.112 | <0.001 |
| Total effect | -0.655 | 0.050 | -0.752, -0.552 | <0.001 | -0.701 | 0.047 | -0.781, -0.598 | <0.001 |
| Proportion mediated | 0.221 |  |  |  | 0.257 |  |  |  |

Note: SE=Standard Error; 95% CI= Confidence Interval

In Model 2, the CFA results showed that anxiety, depression, and uncertainty symptom indicator loaded significantly on the psychosocial distress latent variable, with factor loadings of 0.772, 0.935 and 0.460, respectively. We found that higher psychosocial distress variable indicated by anxiety, depression and uncertainty was significantly associated with lower resilience (β = -0.546, p<0.001). Resilience was in turn associated with greater HRQOL (β = 0.330, p<0.001) (Figure 2). Similar to model 1, psychosocial distress was indirectly associated with HRQOL through resilience (indirect effect: β = -0.180, 95% CI: -0.256, -0.112, p< 0.001; Total effect: β = -0.701, 95% CI: -0.781, -0.598, p< 0.001). Resilience accounted for 25.7% of the total effect of this relationship.

**Figure 2.**
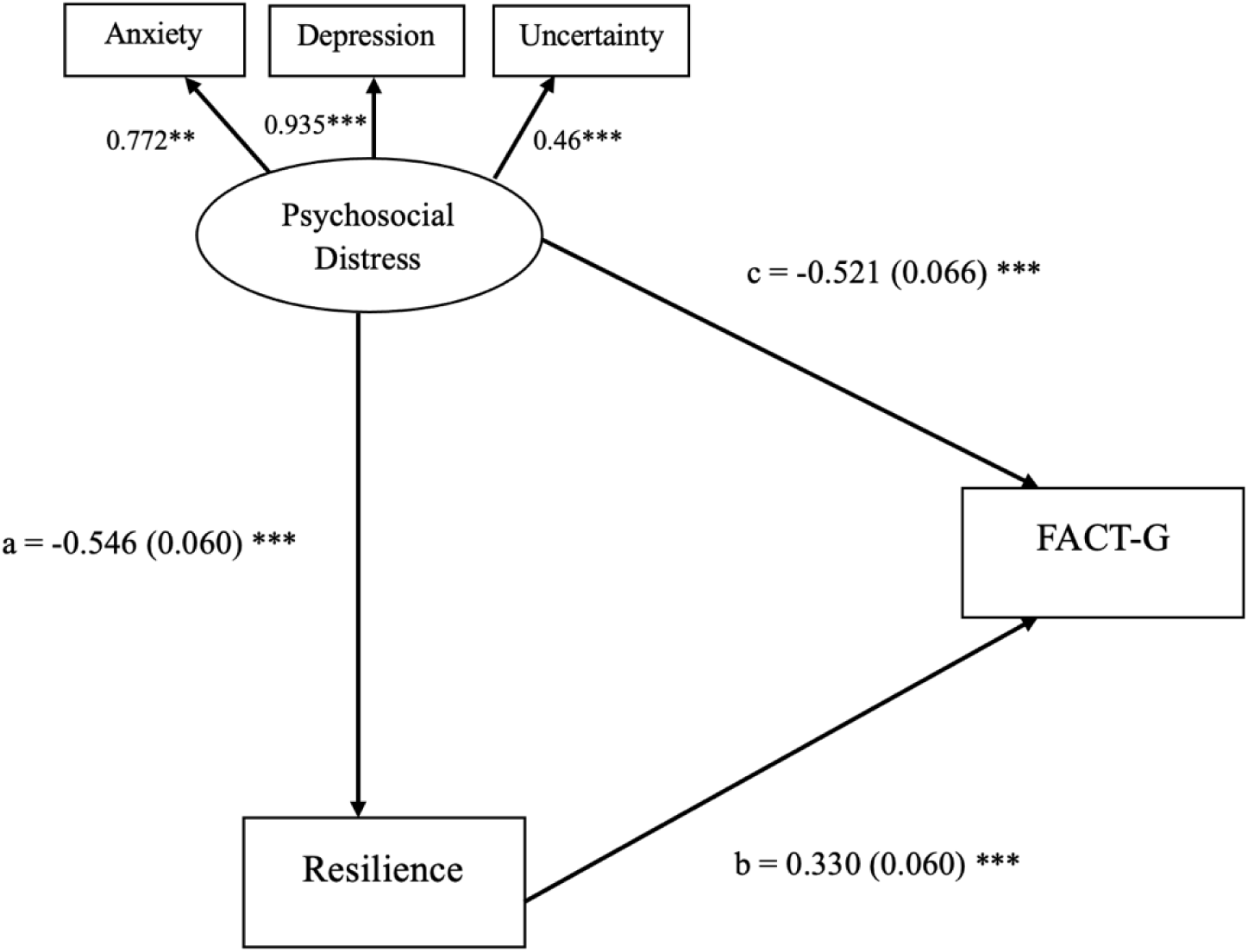
Mediation model of resilience in the relationship between psychological distress and HRQOL (FACT-G) Note: the figure shows factor loading of anxiety, depression and uncertainty on the latent variable psychological distress; a= effects of psychological distress on resilience; b= effects of resilience on FACT-G adjusting for psychological distress and other covariates; c= direct effects of psychological distress on FACT-G adjusting for resilience and covariates. *** p<0.001

In Model 3, we examined the pathways among physical symptom burden, psychosocial distress, resilience and HRQOL (Figure 3). We identified that physical symptom burden was significantly associated with worse psychosocial distress (β = 0.651, p<0.001), which were in turn associated with lower resilience (β = -0.628, p<0.001). Resilience was positively associated with HRQOL (β = 0.334, p<0.001). There was no significant association found between physical symptom burden and resilience (β = 0.108, p= 0.274), as psychosocial distress fully mediated this relationship. We found a significant indirect effect of physical symptom burden on HRQOL through the combination of psychosocial distress and resilience (Indirect effect: β = -0.137, 95% CI: -0.217, -0.078, p<0.001). Table 4 shows the bootstrapping results for each path. Psychosocial distress and resilience together accounted for 47% of the total effect of this relationship.

**Figure 3.**
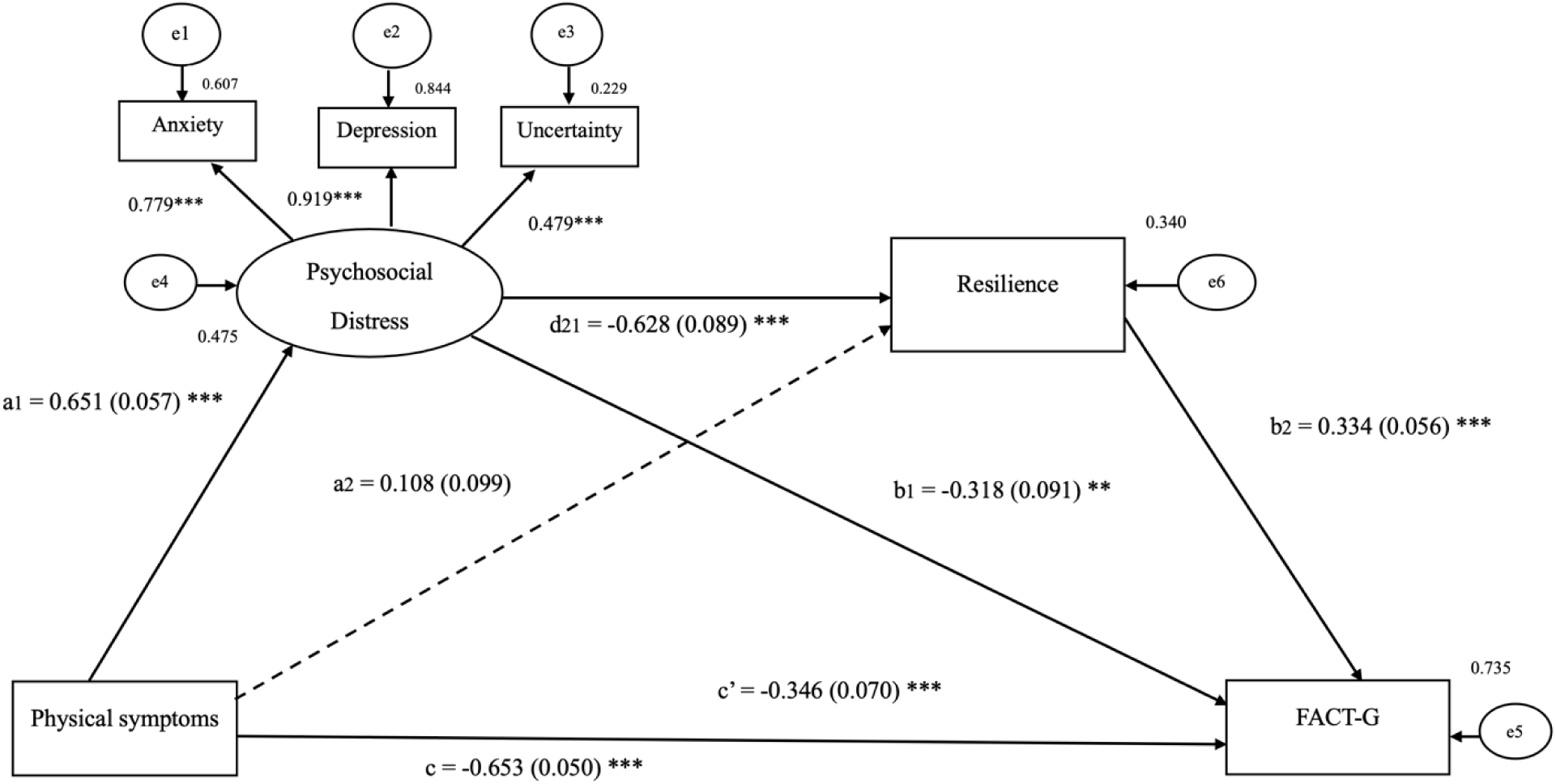
The multiple mediation model of physical symptom burden, psychosocial distress, resilience and HRQOL (FACT-G) Note: the figure includes standardized path coefficients and standard error adjusting for covariates; Goodness-of-fit:X^2^/df = 1.230; RMSEA = 0.033; SRMR = 0.028; CFI = 0.994; and TLI =0.985; R^2^= 0.735; Dash line represents insignificant association. *** p<0.001

**Table 4.**
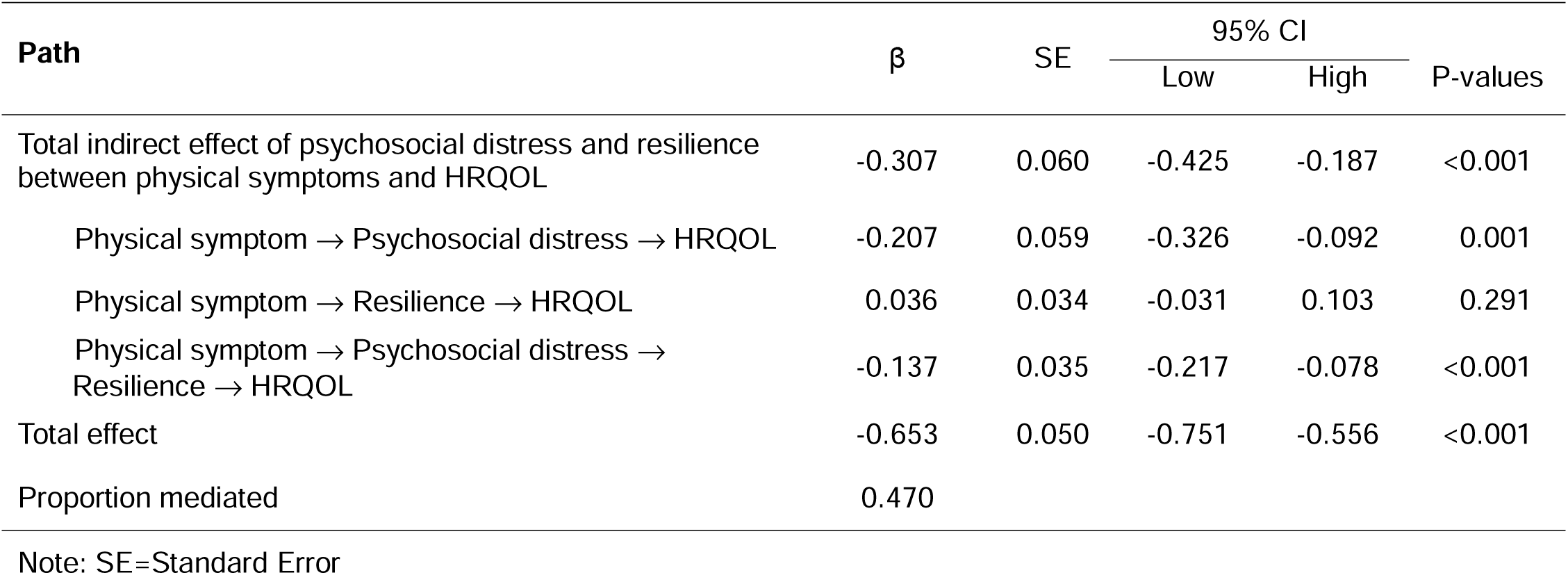
Bootstrapping indirect effect(s) and 95% confidence intervals (CI) for the final structural equation model (Model 3)

### 4.4 Model fit and parameter estimation

Model fit indices for each SEM model are shown in Supplemental Table 2. In Model 1, physical symptoms and resilience together explained 69.4% of the variance in HRQOL (R^2^ = 0.694). In Model 2, psychosocial distress and resilience accounted for 65.1% of the variance in HRQOL, however, the REMSA and SRMR values exceeded 0.08, indicating a suboptimal model fit. Compared with Model 2, Model 3 illustrated a significant improved model, with χ^2^/df = 1.230; RMSEA = 0.033; SRMR = 0.028; CFI = 0.994; and TLI =0.985, indicating an excellent fit of the model, where physical symptoms, psychosocial distress, and resilience collectively explained 73.5% of the variance in HRQOL (R^2^ = 0.735).

## 5. Discussion

In this study, we examined the pathways among physical symptoms, latent psychosocial distress (anxiety, depression, uncertainty), resilience, and HRQOL in women with MBC. Participants in the current study reported moderate levels of physical symptom burden and psychosocial distress, with a mean FACT-G score comparable to prior reports in patients with metastatic cancers [39, 40] but lower than early-stage breast cancer survivors [41] and the general U.S. population [42]. These findings highlighted that, despite advances in contemporary systemic treatment, women with MBC continue to experience substantial symptom burden and poor HRQOL.

Consistent with prior studies, resilience was negatively associated with both physical symptom burden and psychosocial distress and positively associated with HRQOL [23, 24]. Resilience also mediated the relationships of both physical symptom burden and psychosocial distress with HRQOL, suggesting that symptom burden affects quality of life not only directly but also through its impact on women’s resilience. These findings extend prior research demonstrating the role of resilience in mediating effect of cancer related symptom distress on health outcomes [23,24,43,44]. Rather than acting as a trait or independent predictor, resilience operates as a dynamic process through which symptom burden and emotional distress influence HRQOL and overall well-being. A systematic review of resilience and mental outcomes in cancer patients found that patients with high resilience may be better able to mobilize psychological and social resources to cope the disease, and actively engage in self-care, as well as regulate emotional responses, thereby they could maintain optimal quality of life and overall wellbeing [22]. While patients with lower resilience may have limited psychological resources to cope with both physical and psychosocial stressors.

Thes findings were clinically important for MBC population in the contemporary systemic treatment era. Although these newer therapies have extended women’s survivorship, they do not eliminate the persistent physical and psychosocial burden of living with incurable cancer. Unpredictable treatment responses and disease progression required women to continually adapt to the disease over time. Thus, resilience may be an important supportive care target in this context and can be used to support patient’s capacity to maintain functional well-being and HRQOL under persistent stressors.

One key finding was that physical symptom burden was no longer directly associated with resilience after psychosocial distress was included in the final SEM model. This finding indicated that psychosocial distress indicated by anxiety, depression and uncertainty were more closely linked to resilience than physical symptom burden. Although physical symptom burden at some extent contributed to psychosocial distress, severe anxiety and depression can significantly deplete women’s cognitive and emotional resources (e.g., perceived control, self-efficacy, and hope) necessary for resilience and adaptive coping [45–47]. Thus, psychosocial distress may serve an important proximal point for intervention that support resilience among women with MBC.

The unique contribution of this study was that we modeled psychosocial distress as a latent construct indicated by anxiety, depression, and uncertainty. Prior studies usually examined these symptoms as separate predictors of HRQOL [6, 12]; however, in clinical practice, these psychological symptoms frequently co-occur. Women with MBC may simultaneously experience anxiety about disease progression, depression related to functional decline and mortality concerns, and uncertainty about treatment response and prognosis [15]. Modeling these symptoms together allowed us to capture the compound effects of anxiety, depression and uncertainty, which may better reflect the complex emotional responses of living with MBC.

In this study, our participants had higher levels of resilience than other advanced cancer patients reported in prior studies [48, 49]. Several factors may account for this difference. First, our sample was predominantly highly educated, had high rates of private insurance coverage, and may have had better access to healthcare, all of those factors were found to be protective factors of resilience [21]. These structural advantages may provide women with more opportunities to obtain health information, access supportive care and maintain their autonomy. Moreover, many participants were receiving novel systemic therapies, such as CDK4/6 inhibitors or immunotherapy, and had lived with MBC for several years, those factors potentially helped them adapt to illness and develop resiliency.

### 5.1 Implications for Practice and Recommendations for Future Research

Our findings suggested that psychosocial distress, reflected by anxiety, depression and uncertainty collectively had a stronger association with resilience than physical symptom burden alone. In clinical practice, this result underscored the importance of integrating psychosocial assessment, particularly anxiety and depression in supportive care for women with MBC. Screening for anxiety, depression and uncertainty may help identify women whose capacity for resilience is vulnerable and who may be less likely to benefit from resilience intervention.

The results also have implications for resilience-promoting interventions. Although many resilience interventions include stress management components, such as mindfulness mediation practices and muscle relaxation techniques [53], those general stress reduction strategies may be insufficient to address cumulative stressors experienced by MBC patients. Interventions for this population can be strengthened by directly addressing anxiety and depressive symptoms, and uncertainty related to certain events associated with MBC. As addressing psychosocial distress before or alongside resilience interventions may promote women’s ability to engage in self-management, use coping resources, and thus maintain better HRQOL.

In addition, future longitudinal studies are warranted to examine the role of resilience and its associations with symptoms and health outcomes across the disease trajectory. Such studies would help to clarify longitudinal relationships between resilience, symptom burden and health outcomes, and inform optimal timing to deliver resilience-promoting interventions (e.g., at diagnosis, treatment changes, and progression). In addition, qualitative and mixed methods research will be also needed to explore how women define resilience and which strategies they used to build and maintain resilience, and how structural factors promote or constrain their capacity to be resilient.

### 5.2 Limitations of the Study

This study has several limitations. First, we used cross-sectional design, which precludes our ability to examine casual relationships among variables. Second, all data were self-reported, which may contribute to self-selection bias. In addition, participants who were able to complete the questionnaires may be more likely to engage in their care, have better functioning and higher levels of resilience than other MBC patients. Thus, our findings may not be generalizable to other groups of patients with MBC. Finally, most of our participants were White and well-educated women, which limited our ability to examine differences in symptom experiences and health outcomes by race, ethnicity, and socioeconomic status. Future study is warranted to recruit more racially diverse samples to better understand resilience and HRQOL among underrepresented MBC patients.

## 6. Conclusions

Physical symptom burden and psychosocial distress were still the main factors contributing to poor HRQOL in women with MBC despite the advancement in cancer treatment. Resilience could be a modifiable target in the care of this population, and promoting resilience may improve women’s HRQOL and overall well-being. Our study findings also highlighted an opportunity for improving current resilience-promoting interventions for MBC population.

## Data Availability

All data produced in the present study are available upon reasonable request to the authors

## AUTHOR CONTRIBUTIONS

All authors contributed to the study conception and design. Author Y.Z. was responsible for the conception, study design, data analysis, data interpretation and manuscript preparation. S.L.F., S.J., M.L., M.R., D.T., and M.T.K contributed significantly to conception, data interpretation, and preparation of manuscript.

## DISCLOSURES AND ACKNOWLEDGEMENTS

The authors have no relevant financial or non-financial interests to disclose. The authors gratefully acknowledge all the women who participated in this study, Lesley Glenn (CEO/Founder of Project Life), Marlena Murphy (deceased) and Julia Maués (Co-Founder of GRASP Cancer) for their invaluable support with recruitment, and the Breast oncology team at the Yale University Smilow Cancer Center.

## INFORMED CONSENT

Informed consent was obtained from all individual participants included in the study.

