## Supplemental Tables for "Symptom burden, Psychosocial Distress, Resilience and Health-Related Quality of Life in Women with Metastatic Breast Cancer Receiving Contemporary Systemic Therapy"

| **Supplemental Table 1**. Unadjusted and adjusted effects of physical symptom burden, anxiety, depression, and uncertainty on HRQOL | | | | | | | | |
| --- | --- | --- | --- | --- | --- | --- | --- | --- |
|  | **Simple Linear Regression** | | |  | **Multiple Linear Regression** | | | |
|  | β | SE | 95% CI | P | β | SE | 95% CI | P |
| Physical symptom | -0.679 | 0.051 | -0.780, -0.579 | <.001 | -0.674 | 0.063 | -0.799, -0.549 | <.001 |
| *Adjusted R^2^, F(df), P* | 0.458, 176.5 (1), P<.001 | | | | 0.539, 10.63 (25), P<.001 | | | |
| Anxiety | -0.574 | 0.057 | -0.687, -0.462 | <.001 | -0.484 | 0.058 | -0.598, -0.370 | <.001 |
| *Adjusted R^2^, F(df), P* | 0.323, 99.39 (1), P<.001 | | | | 0.460, 8.01 (25), P<.001 | | | |
| Depression | -0.677 | 0.051 | -0.778, -0.576 | <.001 | -0.613 | 0.053 | -0.717, -0.509 | <.001 |
| *Adjusted R^2^, F(df), P* | 0.454, 172.3 (1), P<.001 | | | | 0.571, 11.97 (25), P<.001 | | | |
| Uncertainty | -0.391 | 0.064 | -0.517, -0.265 | <.001 | -0.286 | 0.065 | -0.413, -0.158 | <.001 |
| *Adjusted R^2^, F(df), P* | 0.149, 37.14 (1), P<.001 | | | | 0.323, 4.93 (25), P<.001 | | | |
| Note: SE=Standard Error; 95% CI= Confidence Interval; adjusted for age, race, education, employment status, income, insurance coverage, current treatment(s), palliative care enrollment, and treatment line(s) completed. | | | | | | | | |

| **Supplemental Table 2**. Model fit indices of physical symptom burden, psychological distress and the final mediation structural models | | | | | | | |
| --- | --- | --- | --- | --- | --- | --- | --- |
| **Model** |  | ***X*^2^/*df*** | **RMSEA** | **SRMR** | **CFI** | **TLI** | **R^2^** |
| Model 1 | Physical symptom→resilience→HRQOL | - | 0.000 | 0.000 | 1.000 | 1.000 | 0.694 |
| Model 2 | Psychological distress→resilience→HRQOL | 2.579 | 0.087 | 0.102 | 0.940 | 0.889 | 0.651 |
| Model 3 | Physical symptom→Psychological distress→resilience→HRQOL | 1.230 | 0.033 | 0.028 | 0.994 | 0.985 | 0.735 |
| Note: HRQOL= Health-related quality of life, df=degree of freedom, RMSEA=Root mean square error of approximation, SRMR=Standardized root mean residual, CFI= Comparative Fit Index, TLI=Tucker-Lewis Index | | | | | | | |
